# Routine Early Antibiotic use in SymptOmatic preterm Neonates (REASON): a prospective randomized controlled trial

**DOI:** 10.1101/2020.04.17.20069617

**Authors:** J. Lauren Ruoss, Catalina Bazacliu, Jordan T. Russell, Diomel de la Cruz, Nan Li, Matthew J. Gurka, Stephanie L. Filipp, Richard A. Polin, Eric W. Triplett, Josef Neu

## Abstract

**Objective:** We aim to assess the feasibility of a pragmatic randomized trial of antibiotics vs. no antibiotics in symptomatic premature infants after birth. Most premature infants are exposed to antibiotics after birth without evidence of benefit or harm for this practice. No study to date has attempted to randomize premature infants to antibiotics vs no antibiotics after birth.

**Study Design:** Infants <33 weeks’ gestation admitted to the University of Florida Neonatal Intensive Care Unit were assigned to one of three groups after birth: (A) high risk – antibiotics indicated (B) low risk – antibiotics not indicated (C) eligible for un-blinded randomization (no antibiotics vs antibiotics). The primary outcome is a composite of serious adverse events including (necrotizing enterocolitis, late onset sepsis, bronchopulmonary dysplasia, and death). Odds ratios (and 95% CI’s) were calculated to compare adverse event rates between the two randomized groups.

**Results:** 186 subjects were enrolled (98 infants and 88 mothers) were enrolled over a 2-year period. 56% of infants (n=55) were randomized; 48% of infants randomized to the no antibiotics arm were switched and received antibiotics within the first 48 hours after birth. Serious adverse events were not significantly different between the randomization arms.

**Conclusion:** This is the first prospective randomized trial of antibiotics vs no antibiotics after birth in symptomatic premature infants. The results of this trial establish a framework of feasibility for a larger multicentered trial that is needed to evaluate the risks and benefits of routine antibiotic exposure in premature infants.

## Introduction

Antibiotics are routinely used in symptomatic preterm infants after birth for treatment of presumed early onset sepsis without clear evidence to guide this practice. It has been shown that early use of antibiotics significantly affects the intestinal microbiome and that this effect often persists after discontinuation of the antibiotic.(1-4) Antibiotics alter intestinal ecology inducing gut dysbiosis that may not revert to the original state after the discontinuation of treatment.(1, 5, 6) Clinical evidence has confirmed an association between routine early antibiotic use in preterm infants and increased risk for morbidities. (7-10) Despite concerns for antibiotic exposure a majority of infants born preterm are treated with antibiotics in the first days of life as “standard of care”. (11, 12) The rationale for this practice is based on speculative hypotheses that preterm deliveries may be precipitated by an infection and that it is difficult to distinguish between the respiratory distress of prematurity and neonatal pneumonia or early onset sepsis. In addition, the data shows that culture-confirmed early onset sepsis is actually very low in this population.(13) Thus most infants born <33 weeks’ gestational age are exposed immediately after birth to a potentially dangerous medication for a variable period of time based on dogma rather than scientific evidence.

The origin and evolution of the intestinal microbiome is incompletely elucidated. The characterization of its normal composition and development, identification of factors that disrupt the normal intestinal colonization process, and the consequences of this alteration on a newborn’s future health are important. Understanding the effect of antibiotics on the intestinal microbiome helps in choosing an optimal therapeutic approach and raises questions regarding our current clinical practices. Preliminary studies by our group on fecal samples obtained from preterm infants show that those infants who received antibiotics for more than 7 days had lower diversity indexes at 6 weeks’ post-birth compared to those infants who did not. In addition, emerging evidence from our group (14-17) and other groups (18, 19) shows that necrotizing enterocolitis (NEC) and late onset sepsis (20) are preceded by intestinal dysbiosis characterized by shifts in bacterial genera and this dysbiosis was associated with antibiotic use. (21) Recent literature has also elucidated the connection between this inflammation in the lung and intestinal dysbiosis, the lung – gut axis. (22, 23) The detrimental effect of this widespread antibiotic use may outweigh the potential benefits. Systematic studies that challenge the current therapeutic approach which aim to provide evidence of either benefit or harm of this practice are sorely needed. In addition, understanding the mechanisms though which antibiotic-related intestinal dysbiosis may result in disease would contribute to development of novel therapeutic approaches that would ultimately decrease the burden of prematurity.

The objective of this study was to effectively challenge existing dogma that nearly all preterm infants require at least a short course of intravenous antibiotics because of the risk of early onset sepsis. In this paper, we will present our clinical experience with a pilot study (NCT02784821) of 98 infants born <33 weeks’ gestation that demonstrates the feasibility of a larger adequately powered prospective randomized trial of routine antibiotics vs no antibiotics after birth in symptomatic preterm infants.

## Protocol

### Protocol Development

A two-year internal data review of antibiotic use in <33 weeks’ gestational age infants admitted to the University of Florida was done prior to the protocol design. Approximately 60% of the mothers delivering babies who are born <33 weeks’ gestation received prenatal antibiotics and 80% of these infants received early postnatal antibiotics (<12hours after birth). Our usual postnatal antibiotic regimen (>95%) consisted of intravenously administered ampicillin and gentamicin. According to our unit practice at the time, a majority of infants born <33 weeks’ gestational age regardless of indication for delivery were screened for sepsis and antibiotic therapy was initiated for at least 48 hours after birth. Antibiotics were discontinued at 48 hours if the blood culture was negative, clinical stability was achieved, and complete blood cell count with differential (CBCdiff) and serum C-reactive protein (CRP) were not raising concern for infection. Infants with unstable clinical course or “sicker than expected for gestational age”, with positive blood cultures, or increased inflammatory markers continued the antibiotics at the discretion of the attending physician. Using this information of unit practice patterns, a pragmatic randomized trial of routine parenteral antibiotics versus no antibiotics in symptomatic preterm infants born at <33 weeks’ gestation was designed. The study protocol was presented at multiple neonatology division meetings at the University of Florida in order to obtain a consensus regarding safety of the study, group allocation, switched to receive antibiotics, and routine laboratory evaluation. The study protocol was agreed upon by the division of neonatology prior to commencement of the study.

### Trial Design

The Routine Early Antibiotic use in SymptOmatic preterm Neonates “REASON” is a prospective, un-blinded, randomized pragmatic pilot clinical trial at a single academic center. REASON was designed to evaluate the feasibility and safety of a larger multicenter trial that evaluates the risks and benefits of current antibiotics practice to determine optimal antibiotic use that protects the infants from infection with minimal effect on the microbiome and subsequent adverse outcomes.

### Participants

A goal of 300 infant-mother dyads (150 infants) would be enrolled with 25 infants in group A, 25 infants in group B, and 100 infants in group C over a one year study period. Enrollment goal was changed to 200 infant-mother dyads (100 infants) secondary to admission rate of premature infants <33 weeks’ gestation to the University of Florida Neonatal Intensive Care Unit (NICU). Infants were eligible if born less than 33 weeks’ gestation, admitted to the University of Florida NICU, and without major congenital anomalies that would affect viability. Per the institutional review board (IRB) policy at the University of Florida, mothers are enrolled secondary to the need to access their medical records for perinatal and delivery information. IRB-approved written parental consent for the infant and maternal consent was obtained prenatally or immediately after birth in eligible infants.

### Group assignment and randomization

#### Enrolled infants were assigned to one of three groups

Group A – newborns at high risk for infection, in which antibiotics were indicated, Group B – newborns at low risk for infection, in which antibiotics were not indicated, or Group C – newborns eligible for randomization (Fig. 1, online only and Fig. 2, online only). Group allocation was established by the research team after reviewing the maternal and infant chart and discussing the infant’s status with the bedside clinician. The group allocation was known to the bedside clinician and to the research team. The research team recommended routine laboratory analysis regardless of group allocation, including blood culture and CBCdiff and CRP on admission. The bedside clinician determined whether to obtain the recommended routine laboratory evaluation.

**Figure 1.**
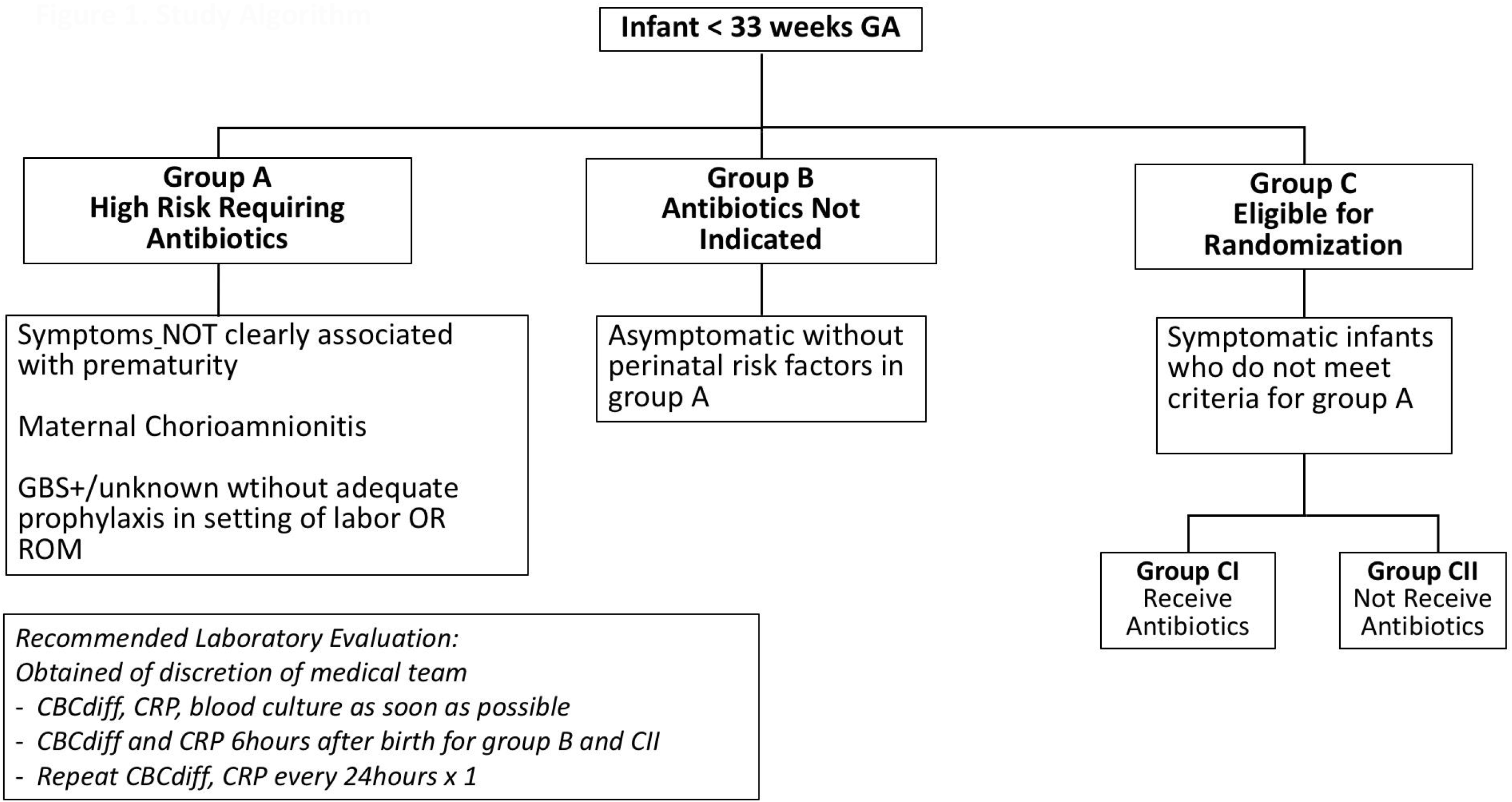
Study Algorithm.

**Figure 2.**
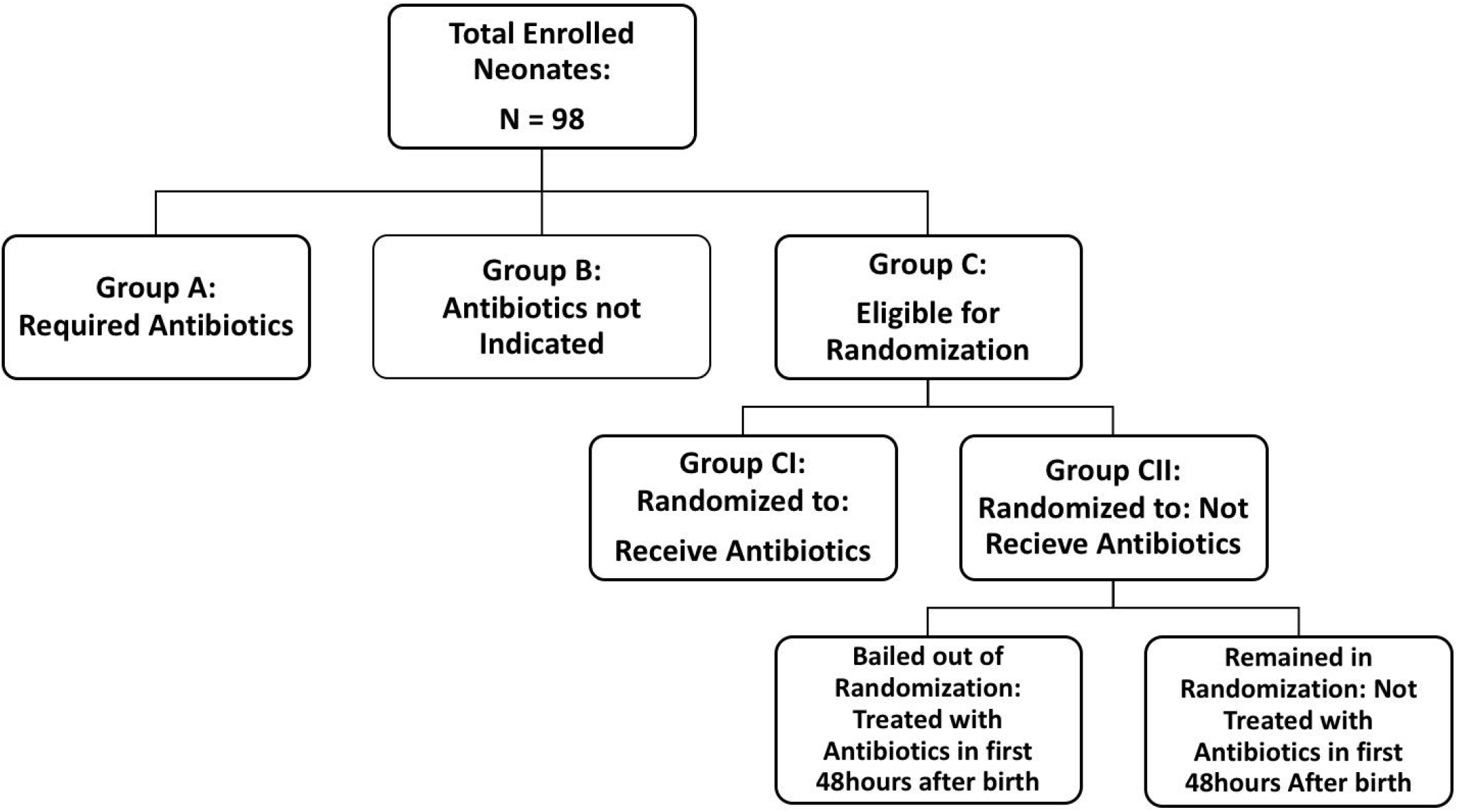
Study Assignment Flow Chart.

#### Group A – high risk - antibiotics indicated

Infants with a higher risk of sepsis were assigned to Group A, in which sepsis screening biomarkers and antibiotic therapy was initiated per the unit practice. Criteria for inclusion in Group A were derived from the Summary of the 2015 NICHD Workshop on Evaluation and Management of Women and Newborns with a Maternal Diagnosis of Chorioamnionitis (24). The criteria for group A included suspected or confirmed chorioamnionitis based on triple I criteria, infants of GBS positive mothers that did not receive adequate prophylaxis, and infants with postnatal symptoms not clearly associated with prematurity. Symptoms not clearly associated with prematurity included but not limited to persistent hypoglycemia, hypotension requiring cardiogenic support, or severe or worsening respiratory failure. This represents about 15% of the patients admitted to our NICU.

#### Group B – low risk - antibiotics not indicated

Infants with very low risk for infection and clinically asymptomatic, showing no significant signs of respiratory distress or having other risk factors based on prenatal and delivery history were enrolled in the second group. This represents about 20% of patients <33 weeks’ gestational age admitted to our NICU.

#### Group C – randomized to receive or not receive antibiotics shortly after birth

Clinically symptomatic premature infants admitted in our NICU who did not meet criteria for Group A were assigned to this group. Infants without symptoms who had sepsis risk factors (chorioamnionitis or GBS positive with inadequate antibiotic prophylaxis) were not eligible for group C. Symptoms were defined as respiratory distress requiring respiratory support, apnea and/or bradycardia, or other factors clearly associated with prematurity. Those were the infants that traditionally were started on pre-emptive antibiotic therapy after birth. The infants in this group were randomized into CI “receive antibiotics” or CII “no antibiotics” arms. Standard open label University of Florida Clinical Research Center methodology was used for randomization. Participants were randomized within one hour of the birth by block randomization using random block sizes of two and four created in SAS and uploaded in REDCap for the use of the study team. The randomization group was then disclosed to the bedside clinician by the research team. The bedside clinician would order the antibiotics if the infant was assigned to the antibiotics group. The intent was that a baby randomized to “receive antibiotics” would be treated for maximum 48 hours. We estimated that approximately 65% of our NICU admissions would qualify for randomization. The main focus of the study is on group C which by randomization will account for other variables including but not limited to mode of delivery, maternal antibiotics, maternal stress.

#### Allowing the group randomized to no antibiotics Group CII, or Group B to receive antibiotics

Infants randomized to no antibiotics or Group B could begin to receive antibiotics at the discretion of the medical team if there was a concern for patient safety. The guidelines or clinical/laboratory criteria used to provide antibiotics was determined by the medical team, not the study. The reason for providing antibiotics to Groups B and CII babies was documented in the clinical chart.

### Risks of intervention and safety mechanisms

Infants assigned to “conventional” treatment of receiving antibiotics immediately after birth if symptomatic (Group A or CI) or not receiving antibiotics when asymptomatic (Group B) were exposed to no additional risk due to study enrollment. Symptomatic infants randomized to “no antibiotics” (Group CII) could have potentially been exposed to higher risk of infection, morbidity, and mortality. To minimize the risks of enrolled infants, the study team recommended obtaining routine laboratory work in all infants. All enrolled infants had blood cultures, CBCdiff, and CRPs drawn shortly after birth and/or at 6 hours after birth, and 24-48hours after birth as per routine care in our NICU. Infants whose blood cultures were positive at any time had intravenous antibiotics initiated per clinical team discretion. The medical care team was allowed to switch any infant randomized to group CII “no antibiotics” or assigned to Group B “antibiotics not indicated” to receive antibiotics, based on their best clinical judgment. The reason for exclusion was documented in the clinical chart.

### Outcomes

The primary outcome was defined prospectively as a composite outcome of late onset sepsis (defined as culture positive infection >48hours after birth), BPD (defined as oxygen requirement at 36 weeks corrected gestation), NEC (defined as Bell’s stage II or greater), and death. Secondary outcomes were defined as early onset sepsis (defined as culture positive infection in the first 48 hours after birth), intraventricular hemorrhage, periventricular leukomalacia, retinopathy of prematurity, and spontaneous intestinal perforation. These outcomes were assessed by review of the medical record at time of discharge from the NICU.

## Study Procedures

### Infant fecal, gastric, and maternal breast milk collection

Infant fecal material and gastric fluid, and maternal breastmilk samples were collected over the course of the NICU hospitalization for each subject. The initial gastric aspirate was obtained if a gastric decompression tube was placed as standard of care and gastric fluid was present at time of birth. An attempt was made to collect the first meconium and then weekly fecal samples were obtained for the duration of hospitalization. A sample of mother’s own milk was obtained in the first week after birth from mothers who choose to pump breastmilk in the first 3 days after birth (Fig. 2, online only).

### Clinical data

Extensive metadata from both mothers and infants were collected from our hospital electronic medical record system and transferred into the electronic database, REDCap®: Research Electronic Data Capture. REDCap is web-based software designed to support clinical data capture and storage. The database was designed to have multiple collection instruments to maintain clinical information for enrolled neonates and their mothers. Data were entered by trained research coordinators on a daily basis throughout the study period and monitored weekly by a statistician for quality checks and safety monitoring.

Covariates from the mother including antepartum antibiotic use/type/duration, length of rupture of membranes, gender, mode of delivery, reason for delivery, pertinent antenatal medications, and placental pathology were recorded. Covariates from the infant included gestational age, feeding information, growth parameters, compete blood counts and inflammatory markers and, antibiotic use/type/duration at any point during the hospitalization.

### Sample Size and Statistical Analysis

Sample size was chosen to ensure sufficiently reliable estimates that provide preliminary evidence that supports our hypotheses and demonstrate feasibility to inform a larger trial. Descriptive statistics, including means and rates, were calculated in this feasibility trial, with corresponding 95% confidence intervals (CI’s) to inform plausible ranges of the feasibility and safety outcomes of interest. Statistical comparisons among groups were made via t-tests and chi-square tests for continuous and categorical data, respectively. Odds ratios (and 95% CI’s) were calculated to compare adverse event rates between the two randomized groups. Data management and descriptive analyses were conducted using SAS 9.4® (Cary, NC).

### Summary of Protocol Changes

The initial study proposal was for 300 infant-mother dyads (150 infants) over a one year period. This was subsequently changed to 200 infant-mother dyads (100 infants) over a two year period based on admission and enrollment rate. The initial study proposal did not include all secondary outcomes that were recorded. These outcomes were added later secondary to noting these morbidities in neonates <33 weeks’ gestation. The initial study proposal did not contain a detailed algorithm for enrollment and allocation after birth to group A, B, or C. After discussions with the neonatology division at the University of Florida a refined algorithm was agreed upon and added to the study protocol prior to the first patient enrolled.

## Results

### Study approval and reviews

The study protocol was submitted on 11/2015, approved by the IRB at University of Florida in Gainesville on 9/2016, and enrollment started on 01/2017 and completed on 01/2019. Enrollment initially was slower than anticipated for several reasons. Over the course of the study there were 9 revisions, including revisions to add personnel to the study, additions of variables included, and amendments to the informed consent. Multiple meetings were held with the IRB secondary to the adverse events including two continuing reviews. There were 46 reportable events during the course of the study. Due to increased mortality and morbidity in infants at the periviable gestation (23-24 weeks), the study was paused for a full IRB board review. The study was resumed given that the incidence of adverse events was similar to the national standard as shown by Vermont Oxford data.

An independent Data Safety and Monitoring Board reviewed all serious adverse events and had two scheduled reviews during enrollment.

### Enrollment and Outcome Data

197 mothers were screened, 124 mothers were consented, and 98 infants and 88 mothers were enrolled. (Fig. 4) Of those consented, 26 infants were excluded (20 screen failure - born after 33 weeks’ gestation, 1 mother declined after consenting, 1 infant died in the delivery room, and 4 infants were excluded because the study team was not called after birth). 98 infants were ultimately enrolled with 32 infants in group A, 11 infants in group B, and 28 infants randomized to group CI (antibiotics) and 27 infants randomized to group CII (no antibiotics). (Table 1) Infants were analyzed according to intention to treat in the assigned group. The rate of infants moving from randomized to no antibiotics (group CII) to being switched to receive antibiotics within the first 48 hours after birth was 48%. Antibiotics were continued beyond 48 hours in 43% of infants assigned to group CI. (Table 2) The trial was ended after the two-year period when a sufficient number of neonates was enrolled.

**Table 1.** Baseline Characteristics.

|  |  | All Study Groups |  |  |  | Randomized Neonates |  |  |
| --- | --- | --- | --- | --- | --- | --- | --- | --- |
|  | Overall | Group B:<br>No Abx Req. | Group C:<br>Randomize | Group A:<br>Abx Req. | p-value† | Group CI:<br>Abx | Group CII:<br>No Abx | p-value† |
| (n) | 98 | 11 | 55 | 32 |  | 28 | 27 |  |
| Male | 50 (51.0) | 4 (36.4) | 30 (54.6) | 16 (50.0) | 0.5400 | 13 (46.4) | 17 (63.0) | 0.2183 |
| C-Section Delivery | 58 (59.8)<sup>1</sup> | 3 (27.3) | 36 (66.7)<sup>1</sup> | 19 (59.4) | 0.0523 | 17 (63.0)<sup>1</sup> | 19 (70.4) | 0.5637 |
| Singleton | 75 (76.5) | 10 (90.9) | 41 (74.6) | 24 (75.0) | 0.4895 | 20 (71.4) | 21 (77.8) | 0.5889 |
| Gestational Age at Birth | 29.1 ± 2.8<br>[23.1, 32.9] | 32.1 ± 1.0<br>[29.3, 32.9] | 29.0 ± 2.6<br>[23.7, 32.7] | 28.2 ± 2.9<br>[23.1, 32.9] | 0.0002 | 29.2 ± 2.5<br>[23.7, 32.7] | 28.8 ± 2.8<br>[23.7, 32.7] | 0.5945 |
| Weight (g) at Birth | 1240 ± 479<br>[490, 2770] | 1900 ± 417<br>[1110, 2770] | 1167 ± 407<br>[525, 2425] | 1138 ± 446<br>[490, 2132] | <0.0001 | 1234 ± 424<br>[525, 2425] | 1098 ± 384<br>[605, 2116] | 0.2160 |
| LOS | 56.4 ± 37.1<br>[0.0, 184.0] | 26.5 ± 19.7<br>[9.0, 81.0] | 57.7 ± 35.2<br>[1.0, 147.0] | 64.5 ± 40.5<br>[0.0, 184.0] | 0.0112 | 53.9 ± 30.0<br>[13.0, 134.0] | 61.6 ± 40.1<br>[1.0, 147.0] | 0.4226 |

†Presented: n(%) for categorical data; Chi-Square Test, and means ± STD [min, max]; ANOVA for continuous data
<sup>1</sup>n missing = 1; did not obtain consent to collect maternal data

**Table 2.**
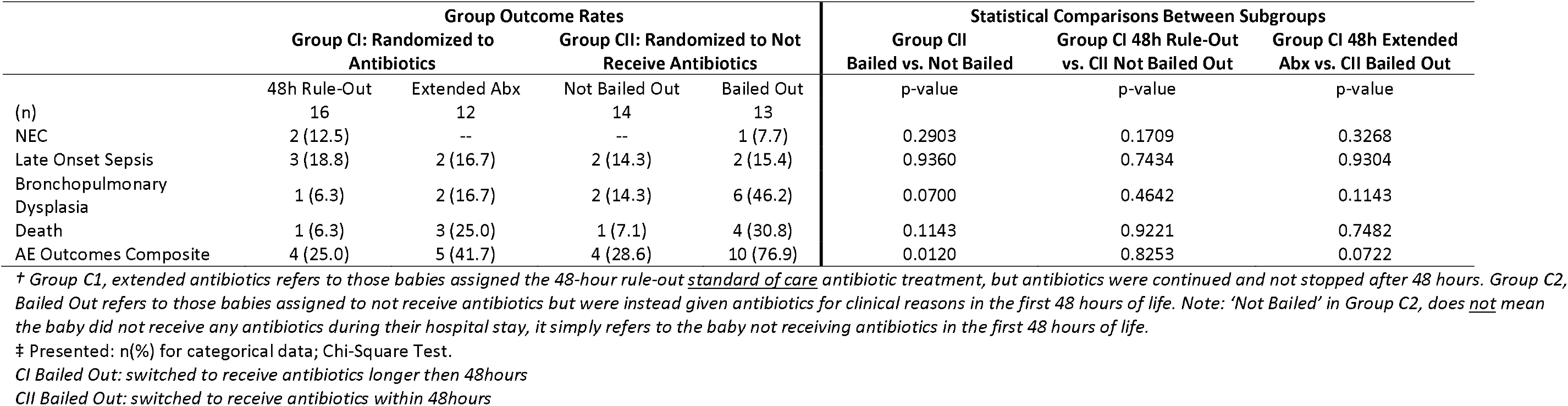
Adverse Events Outcomes Within Randomized Babies.

**Figure 3.**
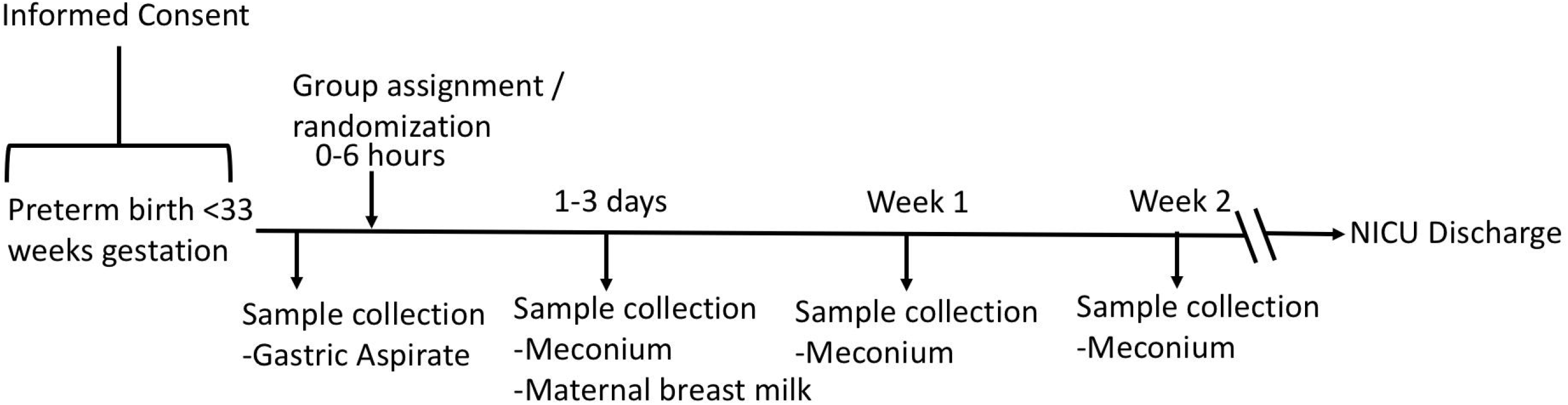
Study Timeline. Collection Timeline.

**Figure 4.**
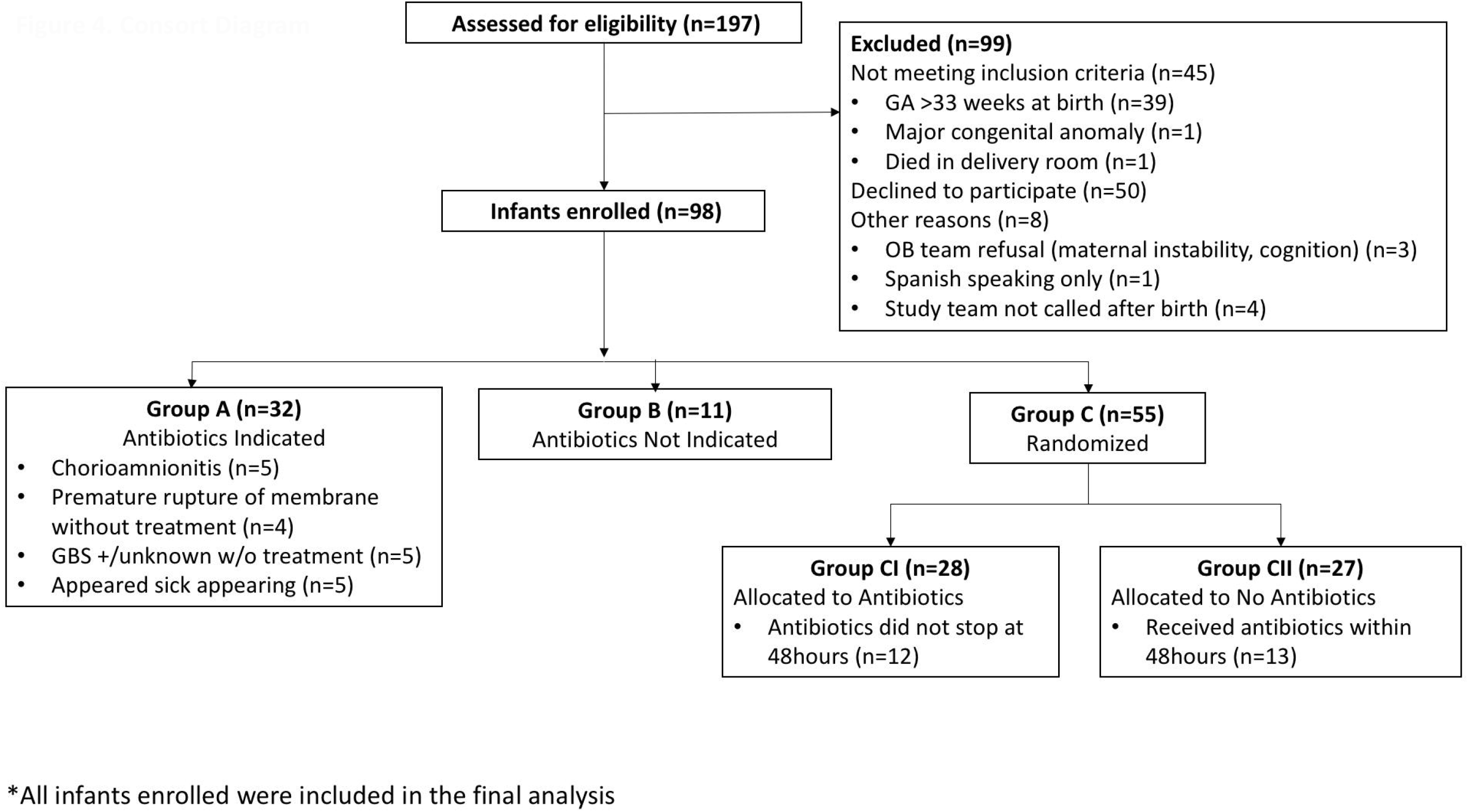
Consort Diagram.

### Switched from randomized to no antibiotic to receiving antibiotics

Thirteen infants (48%) were switched from group CII (no antibiotics) to receiving antibiotics. Seven infants were switched to antibiotics in group CII for laboratory abnormalities: one – elevated white blood cell count, three – elevated CRP, and three for elevated CRP along with documentation of increased fraction of inspired oxygen requirement without other changes in clinical status. Six infants were switched to antibiotics in group CII for clinical deterioration: two received chest compressions, one spontaneous intestinal perforation, one septic shock, and one hemorrhagic shock requiring continuous cardiogenic support. Only one infant had culture positive early onset sepsis in the study and this infant was in group C randomized to no antibiotics and switched to antibiotics at one hour after birth. Three infants were treated for culture for negative early onset sepsis.

### Sample collection

A total of 693 stool samples (median of 7 samples per patient) and 20 gastric aspirates were collected from 91 infants. Mother’s own milk samples were collected for 78 infants. Stool samples could not be collected from 7 infants due to early mortality.

### Safety and Sudden Adverse Events

Individual adverse events were not significantly different between enrollment groups. (Table 3) A composite adverse event outcome encompassing NEC, late onset sepsis, bronchopulmonary dysplasia and death was significantly different. As expected based on the design of the study, the rate of individual and composite of serious adverse events was lower in the infants assigned to group B than in groups A and C. The composite outcome between randomized groups CII vs CI was different (51.9% vs 32.1%; OR=2.27 95% CI= [0.76, 6.80]), though not statistically significant in this small pilot study. There was a notable difference in the rate of BPD (29.6% n=8 vs. 10.7% - n=3, vs for group CII vs CI respectively; OR=3.51, 95% CI= [0.82, 15.03]) among randomized babies, though not statistically significant. (Table 3)

**Table 3.**
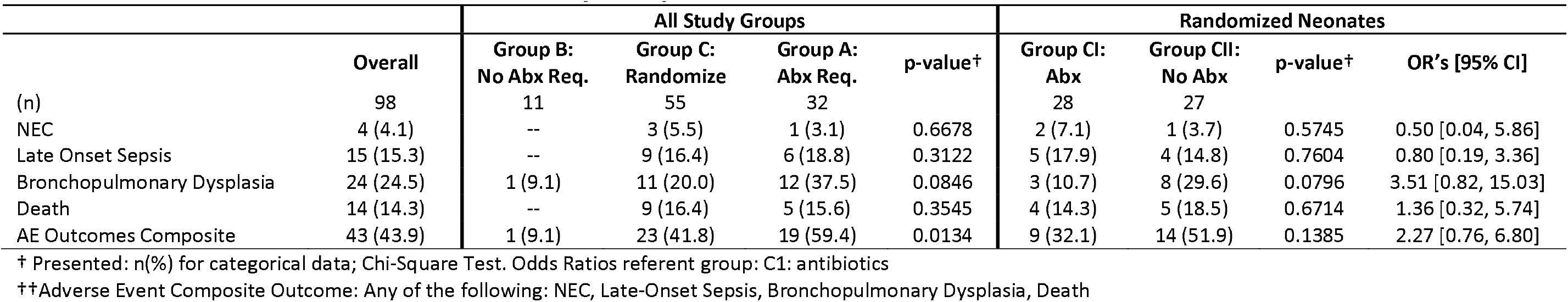
Adverse Events Outcomes Across Study Groups.

Five (15.6%) infants enrolled in Group A died and 9 infants (16.4%) of infants in group C died. One infant in the study had early onset sepsis and that infant died in the first 24hours after birth from a bacteria resistant to standard antibiotic treatment. There was no statistically significant difference between the numbers of deaths in infants in group CI (14.3%) vs group CII (18.5%). All infants assigned to group B survived to discharge. Infants in group B had a shorter length of stay in comparison with all other groups, concordant with an older gestation. Randomization to receive or not antibiotics did not significantly impact the length of hospitalization. Placement into group C mitigates degree of illness as infants placed into group B were asymptomatic and infant’s placed into group A either had sepsis risk factors as previously defined or had a severity of illness that was not expected for estimated gestation.

### Status of study

All infants have completed enrollment. Clinical data was reviewed for quality assurance. All meconium, stool, gastric, and breastmilk samples have been collected and final analysis of microbiome, fecal inflammatory markers, and metabolomics is expected to be soon completed.

## Discussion

We have shown that a study that challenges the current practice of routine antibiotics in premature symptomatic infants is feasible and can be done in a manner that causes no harm. To our knowledge, this is the first study to randomize symptomatic premature infants to receive early parenteral antibiotics versus no antibiotics after birth. While this paper demonstrates feasibility of such a trial, a multicenter study is needed to confirm our findings of no harm and to change standard of care. Our experience with this study provides important points that will guide us in the next trial. Two important aspects for an expansion of this trial include the difficulties experienced with the infants at periviable gestational ages and the lack of increased adverse events in infants randomized to antibiotics in comparison to infants not randomized to antibiotics. These points are described in further detail below and may be used to guide further trial design.

Enrollment was slower than originally expected. This was in part due to multiple reviews by the IRB for serious adverse events. A majority of these reviews were triggered by adverse events unrelated to the study, that were suffered by infants at the periviable gestation (23-24 weeks estimate gestation) who were assigned to Group A. Expansion of this trial and future trials evaluating antibiotic use after birth should consider excluding infants with high likelihood of early morbidity and mortality such as neonates at periviable gestation and infants with high illness severity scores. Newly developing scoring systems like nSOFA may better assist in the population selection process.(25) The study was completed in January 2019 (two years after commencement) after enrollment of 98 infants. While the goal enrollment was not met, this number of subjects was determined to be sufficient to determine feasibility. Given that infants at the periviable gestation had increased morbidity and mortality, it would be prudent in further trial designs to exclude infants <25 weeks gestation.

In this pilot trial, statistically significant increases in adverse outcomes were not found among infants assigned to no antibiotic use in the first 48 hours after birth (32.1% in CI and 51.9% in CII). The difference in composite outcomes between groups CI and CII is most affected by the different rates in BPD. BPD is a multifactorial disease and the difference in rates between group CI and group CII is likely not due to antibiotic group assignment. Antibiotics have been proposed to decrease inflammation and subsequently decrease ventilator requirements, which would decrease the rate of BPD. However, new studies are investigating the connection between airway and intestinal dysbiosis with abnormal inflammation that alters lung development, subsequently increasing the risk of BPD. (22, 23) In our study, 8 infants in the no antibiotics group developed BPD however, 6 of those infants were switched to antibiotics and received antibiotics in the 48 hours after birth. Based on the small numbers it does not appear that withholding antibiotics in the first 48 hours led to BPD. However, due to the small numbers of infants with BPD (3 in group CI and 8 in group CII), we are unable to conclude what drove the difference in rates of BPD between the group C arms. If adverse events are to be a primary outcome in the larger multicenter study, then a non-inferiority trial will be in order (Table 2). The safety of randomizing premature infants to antibiotics vs no antibiotics has been a major concern in neonatology. Although we are not claiming safety, our pilot study’s pragmatic design showed no initial evidence of harm. It allowed the primary physicians to provide antibiotics based on their clinical judgment. The majority of these were due to abnormal laboratory findings or acute events such as tension pneumothorax leading to chest compressions or spontaneous intestinal perforation. Only one infant in the study had culture positive early onset sepsis. This infant was randomized to no antibiotics (group CII) however was administered antibiotics at one hour after birth secondary to clinical status change and laboratory evaluation as outlined in study algorithm. Standard antibiotics were initiated (ampicillin and gentamicin) however due to continued clinical deterioration the antibiotics were broadened. The infant was started on standard antibiotics by one hour after birth, which is within our NICU’s practice, and serial exams and early laboratory evaluation accurately detected the infant’s severity of illness. The infant died from early onset sepsis with a bacterial strain resistant to standard antibiotics. This infant’s case was reviewed by the data safety monitor board, the neonatology division, and IRB at the at the University of Florida which determined the death was not study related as the infant was changed to antibiotics in a timely manner. The low rate of early onset sepsis observed in this cohort is similar to other previously published studies and reinforced the goal of challenging the practice of routine antibiotic use to prevent early onset sepsis. (13, 26)

As there is no consensus on a laboratory marker suggestive of infection, clinical providers used their own judgment on antibiotic use. While this approach may have encouraged participation by neonatologists in the study, it also created an inconsistent approach which is a limitation of this trial. Expansion of this trial and future randomized trials evaluating on this topic should consider blinding the treatment group to improve consistency and potentially decrease the rate of switching infants from no antibiotic use to receiving antibiotics. During the course of this study, a decrease in the use of antibiotics in our NICU occurred in infants with birth weight lower than 1250g, where overall duration of empiric antibiotic use decreased by 1.96 days between 2012 and 2018. The incidence of patients who received an empiric treatment course longer than 5 days also decreased from 58.8% to 23.3% in the same time period. The decrease in antibiotics over the past few years is consistent with the findings of Greenberg et al. (12). This trend of decreasing antibiotic utilization in premature infant care suggests a growing concern with unnecessary antibiotic use, but the overall use remains very high, and demonstrates that a multicenter trial evaluating the antibiotic in neonatal care is needed.

## Conclusions

Early antibiotic use in symptomatic premature infants to decrease early onset sepsis is based on dogma and this practice may be unnecessary. In this paper, we demonstrate that a randomized trial of early antibiotics vs no antibiotics in a neonatal care unit is feasible. Our experiences provide guidance for larger randomized trials, which in turn may improve clinical care of preterm infants.

## Data Availability

Data will be made available upon request.

## Abbreviations

NEC: necrotizing enterocolitis
BPD: bronchopulmonary dysplasia
CRP: serum C-reactive protein
CBCdiff: complete white blood cell count with differential
NICU: Neonatal Intensive Care Unit
IRB: Institutional Review Board

## Acknowledgements

for the Data Safety Monitoring Board: Susmita Datta, PhD - Biostatistician and Epidemiologist at University of Florida, Michael Cotton, M.D. - Neonataologist at Duke University School of Medicine, William Benitz, M.D. - Neonatologist at Lucile Packard Children’s Hospital, Stanford, Robert Lawrence, M.D. - Pediatric Infectious Disease specialist at University of Florida

## References

1. Dethlefsen L, Relman DA. Incomplete recovery and individualized responses of the human distal gut microbiota to repeated antibiotic perturbation. Proc Natl Acad Sci U S A. 2011;108 Suppl 1:4554–61.

2. Greenwood C, Morrow AL, Lagomarcino AJ, Altaye M, Taft DH, Yu Z, et al. Early Empiric Antibiotic Use in Preterm Infants Is Associated with Lower Bacterial Diversity and Higher Relative Abundance of Enterobacter. J Pediatr. 2014.

3. Arboleya S, Sanchez B, Milani C, Duranti S, Solis G, Fernandez N, et al. Intestinal microbiota development in preterm neonates and effect of perinatal antibiotics. J Pediatr. 2015;166:538–44.

4. DiGiulio DB. Prematurity and perinatal antibiotics: a tale of two factors influencing development of the neonatal gut microbiota. J Pediatr. 2015;166:515–7.

5. Dethlefsen L, Huse S, Sogin ML, Relman DA. The pervasive effects of an antibiotic on the human gut microbiota, as revealed by deep 16S rRNA sequencing. PLoS Biol. 2008;6:e280.

6. Fouhy F, Guinane CM, Hussey S, Wall R, Ryan CA, Dempsey EM, et al. High-throughput sequencing reveals the incomplete, short-term recovery of infant gut microbiota following parenteral antibiotic treatment with ampicillin and gentamicin. Antimicrob Agents Chemother. 2012;56:5811–20.

7. Alexander VN, Northrup V, Bizzarro MJ. Antibiotic exposure in the newborn intensive care unit and the risk of necrotizing enterocolitis. J Pediatr. 2011;159:392–7.

8. Cotten CM, Taylor S, Stoll B, Goldberg RN, Hansen NI, Sanchez PJ, et al. Prolonged duration of initial empirical antibiotic treatment is associated with increased rates of necrotizing enterocolitis and death for extremely low birth weight infants. Pediatrics. 2009;123:58–66.

9. Abdel Ghany EA, Ali AA. Empirical antibiotic treatment and the risk of necrotizing enterocolitis and death in very low birth weight neonates. Ann Saudi Med. 2012;32:521–6.

10. Greenwood C, Morrow AL, Lagomarcino AJ, Altaye M, Taft DH, Yu Z, et al. Early empiric antibiotic use in preterm infants is associated with lower bacterial diversity and higher relative abundance of Enterobacter. J Pediatr. 2014;165:23–9.

11. Schulman J, Dimand RJ, Lee HC, Duenas GV, Bennett MV, Gould JB. Neonatal intensive care unit antibiotic use. Pediatrics. 2015;135:826–33.

12. Greenberg RG, Chowdhury D, Hansen NI, Smith PB, Stoll BJ, Sanchez PJ, et al. Prolonged duration of early antibiotic therapy in extremely premature infants. Pediatr Res. 2019;85:994–1000.

13. Stoll BJ, Gordon T, Korones SB, Shankaran S, Tyson JE, Bauer CR, et al. Early-onset sepsis in very low birth weight neonates: a report from the National Institute of Child Health and Human Development Neonatal Research Network. J Pediatr. 1996;129:72–80.

14. Torrazza RM, Ukhanova M, Wang X, Sharma R, Hudak ML, Neu J, et al. Intestinal microbial ecology and environmental factors affecting necrotizing enterocolitis. PLoS One. 2013;8:e83304.

15. Mai V, Torrazza RM, Ukhanova M, Wang X, Sun Y, Li N, et al. Distortions in development of intestinal microbiota associated with late onset sepsis in preterm infants. PLoS One. 2013;8:e52876.

16. Mai V, Young CM, Ukhanova M, Wang X, Sun Y, Casella G, et al. Fecal microbiota in premature infants prior to necrotizing enterocolitis. PLoS One. 2011;6:e20647.

17. Pammi M, Cope J, Tarr PI, Warner BB, Morrow AL, Mai V, et al. Intestinal dysbiosis in preterm infants preceding necrotizing enterocolitis: a systematic review and meta-analysis. Microbiome. 2017;5:31.

18. Wang Y, Hoenig JD, Malin KJ, Qamar S, Petrof EO, Sun J, et al. 16S rRNA gene-based analysis of fecal microbiota from preterm infants with and without necrotizing enterocolitis. ISME J. 2009;3:944–54.

19. Carl MA, Ndao IM, Springman AC, Manning SD, Johnson JR, Johnston BD, et al. Sepsis from the gut: the enteric habitat of bacteria that cause late-onset neonatal bloodstream infections. Clin Infect Dis. 2014;58:1211–8.

20. Madan JC, Farzan SF, Hibberd PL, Karagas MR. Normal neonatal microbiome variation in relation to environmental factors, infection and allergy. Curr Opin Pediatr. 2012;24:753–9.

21. Neu J, Pammi M. Pathogenesis of NEC: Impact of an altered intestinal microbiome. Semin Perinatol. 2017;41:29–35.

22. Casado F, Morty RE. The emergence of preclinical studies on the role of the microbiome in lung development and experimental animal models of bronchopulmonary dysplasia. Am J Physiol Lung Cell Mol Physiol. 2020.

23. Tirone C, Pezza L, Paladini A, Tana M, Aurilia C, Lio A, et al. Gut and Lung Microbiota in Preterm Infants: Immunological Modulation and Implication in Neonatal Outcomes. Front Immunol. 2019;10:2910.

24. Higgins RD, Saade G, Polin RA, Grobman WA, Buhimschi IA, Watterberg K, et al. Evaluation and Management of Women and Newborns With a Maternal Diagnosis of Chorioamnionitis: Summary of a Workshop. Obstet Gynecol. 2016;127:426–36.

25. Wynn JL, Polin RA. A neonatal sequential organ failure assessment score predicts mortality to late-onset sepsis in preterm very low birth weight infants. Pediatr Res. 2019.

26. Kuzniewicz MW, Walsh EM, Li S, Fischer A, Escobar GJ. Development and Implementation of an Early-Onset Sepsis Calculator to Guide Antibiotic Management in Late Preterm and Term Neonates. Jt Comm J Qual Patient Saf. 2016;42:232–9.

